# Challenges in control of COVID-19: short doubling times and long delay to effect of interventions

**DOI:** 10.1101/2020.04.12.20059972

**Authors:** Lorenzo Pellis, Francesca Scarabel, Helena B. Stage, Christopher E. Overton, Lauren H. K. Chappell, Katrina A. Lythgoe, Elizabeth Fearon, Emma Bennett, Jacob Curran-Sebastian, Rajenki Das, Martyn Fyles, Hugo Lewkowicz, Xiaoxi Pang, Bindu Vekaria, Luke Webb, Thomas A. House, Ian Hall

## Abstract

The unconstrained growth rate of COVID-19 is crucial for measuring the impact of interventions, assessing worst-case scenarios, and calibrating mathematical models for policy planning. However, robust estimates are limited, with scientific focus on the time-insensitive basic reproduction number *R*0. Using multiple countries, data streams and methods, we consistently estimate that European COVID-19 cases doubled every three days when unconstrained, with the impact of physical distancing interventions typically seen about nine days after implementation, during which time cases grew eight-fold. The combination of fast growth and long detection delays explains the struggle in countries’ response better than large values of *R*0 alone, and warns against relaxing physical distancing measures too quickly. Testing and tracing are fundamental in shortening such delays, thus preventing cases from escalating unnoticed.

## 1. Introduction

In December 2019, a cluster of unexplained pneumonia cases in Wuhan, the capital of Hubei province in the People’s Republic of China, rapidly progressed into a large-scale outbreak, and a global pandemic by 11 March 2020, as declared by the World Health Organisation (1). The disease caused by this highly contagious infection has since been named COVID-19, and is caused by a single-stranded RNA coronavirus (SARS-CoV-2) similar to the pathogen responsible for severe acute respiratory syndrome (SARS) and Middle East respiratory syndrome (MERS).

Planning interventions, in particular physical distancing, usually relies on estimates of the basic reproduction number *R*_0_. This is defined as the average number of new infections generated by a single infected person in a fully susceptible population without control in place, and determines the portion of transmission that needs to be prevented to avoid spread (2). The impact of interventions is often quantified by the difference between *R*_0_ and the effective reproduction number *R_e_* (sometimes denoted *R*_t_, or simply *R*), *R*_0_’s counterpart in the presence of control measures and/or population acquired immunity.

Early published estimates of COVID-19 *R*_0_ were extremely variable, ranging from 1.4 to 6.49 (3, 4), with most official sources reporting values in the range of 2-3 (5-8). However, these latter values mostly derive from early studies of the epidemic in Wuhan (9-12) or the Diamond Princess Cruise ship (13), and so are subject to important limitations, including small amounts of data, arbitrary assumptions on epidemiological parameters (for instance, assumed serial/generation intervals from SARS and MERS), uncertain or biased reporting of early cases, and the uniqueness of the specific settings in which they occurred. For these limitations, continuous effort should be devoted to understanding the discrepancies in published values, and official ranges of *R*_0_ should be continuously updated with refined estimates both from China (14, 15), and other nations (16-19).

Nevertheless, over-reliance on *R*_0_ should be avoided. First, *R*_0_ lacks temporal information related to the speed of epidemic growth, and hence the optimal timing of interventions (20, 21). Second, estimates of *R*_0_ can vary considerably – and in particular do so for COVID-19 – as a reflection of both genuine differences in geography and settings, but also how it is calculated: *R*_0_ is typically indirectly derived through mathematical models, with values varying depending on model structure and estimates of (or assumptions on) parameter values, even when the same data are used for model fits.

We argue that the real-time growth rate and the delay between infection and case detection are often more informative than precise estimates of *R*_0_ for initiating interventions (22, 23): since doubling times and delays can be inferred directly from incidence and line-list data, sophisticated models are not required to infer when action is urgent. Similarly, as interventions are lifted, the growth rate can offer a more direct indicator of the current trend in cases than *R_e_*, whilst understanding delays in detection is vital to inform the appropriate intervals between subsequent relaxations.

While numerous estimates of *R*_0_ for COVID-19 exist, real-time growth rates are much less prominent in the literature and, where published, lack robustness or are restricted to single-country analyses on a single dataset (16, 24). Even with interventions in place, estimates of unconstrained growth remain essential for pandemic response, in particular to calibrate the dynamics of mathematical models which are then used to assess the impact of interventions, plan worst-case scenarios and explore short- and long-term forecasts.

## 2. Results

Here, we provide robust estimates of the initial growth rate in multiple European countries, obtained with different methods. We then estimate the incubation period and the distribution of times from symptom onset to hospitalisation under multiple scenarios and in different settings, and compare them with other results from the literature. Our results show that, in an unconstrained epidemic, cases can grow 8-fold before the effect of any intervention becomes visible. Observation of the initial outbreak in the UK and Italy provides further support to the highlighted delays from infection to detection.

To estimate the growth rate, we focussed on the number of confirmed cases in the most affected European countries (Figure 1A), as reported by the WHO (25) on 31 March 2020. The analysis accounts for changes in reporting by day of the week. To avoid relying only on case confirmation, which could be affected by numerous biases, we also estimated the growth rate from hospital and intensive care unit (ICU) bed occupancy and deaths in Italy (Figure 1B; 26). To ensure generalisability of results, we performed another analysis on a larger set of European countries (Figure 2). With the exception of a few countries that are known to have changed the testing policy over time or whose data appear visibly noisy, we consistently found doubling times of about 3 days, that appear to be sustained before mitigation interventions are put in place. These values are significantly shorter than early estimates from China (9, 12). For robustness, we have used two different methods: semi-parametric and generalised linear (details in Materials and Methods). Unsurprisingly, the results differ in terms of their confidence intervals, but the conclusions are similar.

**Fig. 1.**
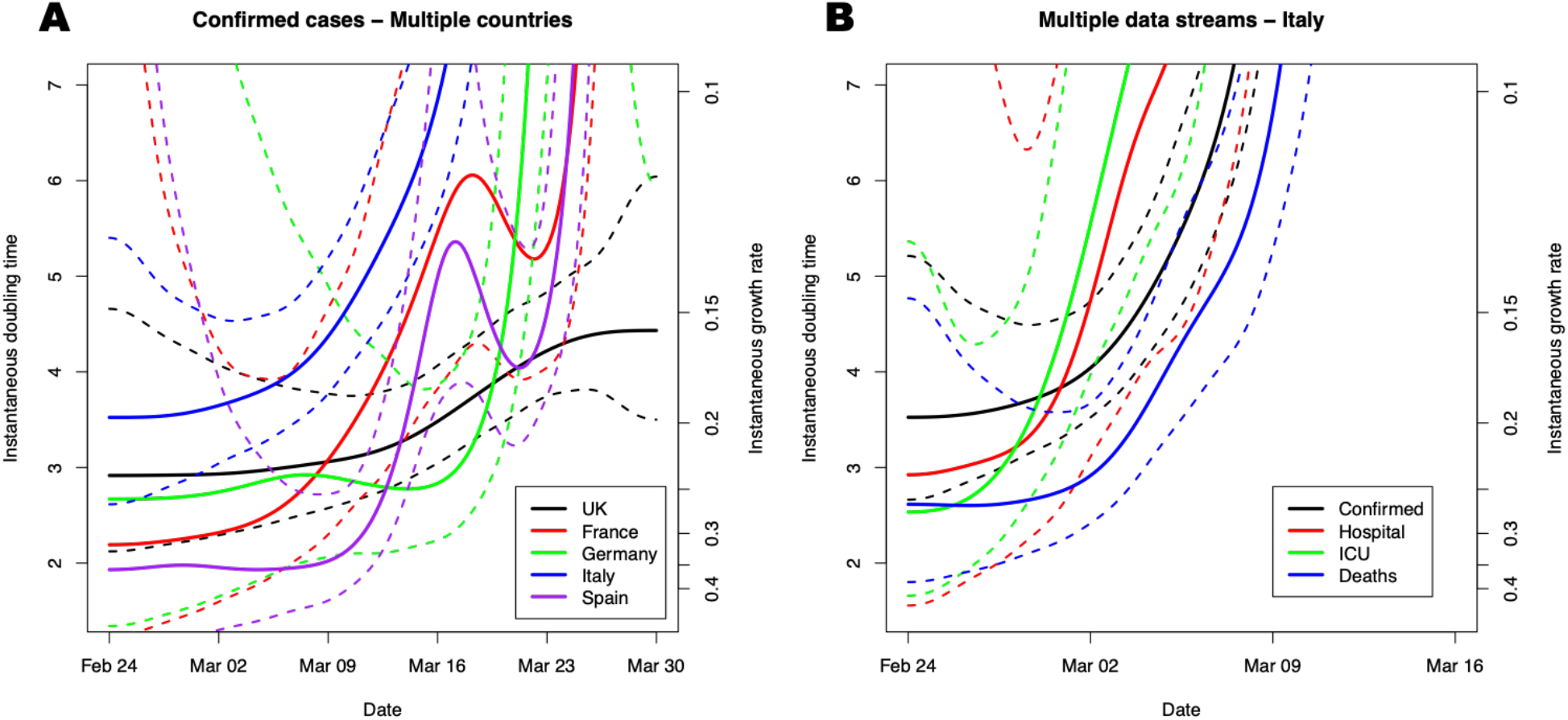
Unconstrained doubling time of about 3 days from multiple countries and data streams. Instantaneous doubling time (left axis) and growth rate (right axis), with 95% confidence intervals (CIs, dashed) obtained by fitting a Generalised Additive Model with quasi-Poisson Family and canonical link to data, adjusted by day-of-week fixed effect (see Materials and Methods), to (A) daily confirmed cases of the five largest European epidemics by the end of March and (B) different surveillance data streams within Italy (daily confirmed cases and deaths, and hospital and ICU daily counts obtained from prevalence).

**Fig. 2.**
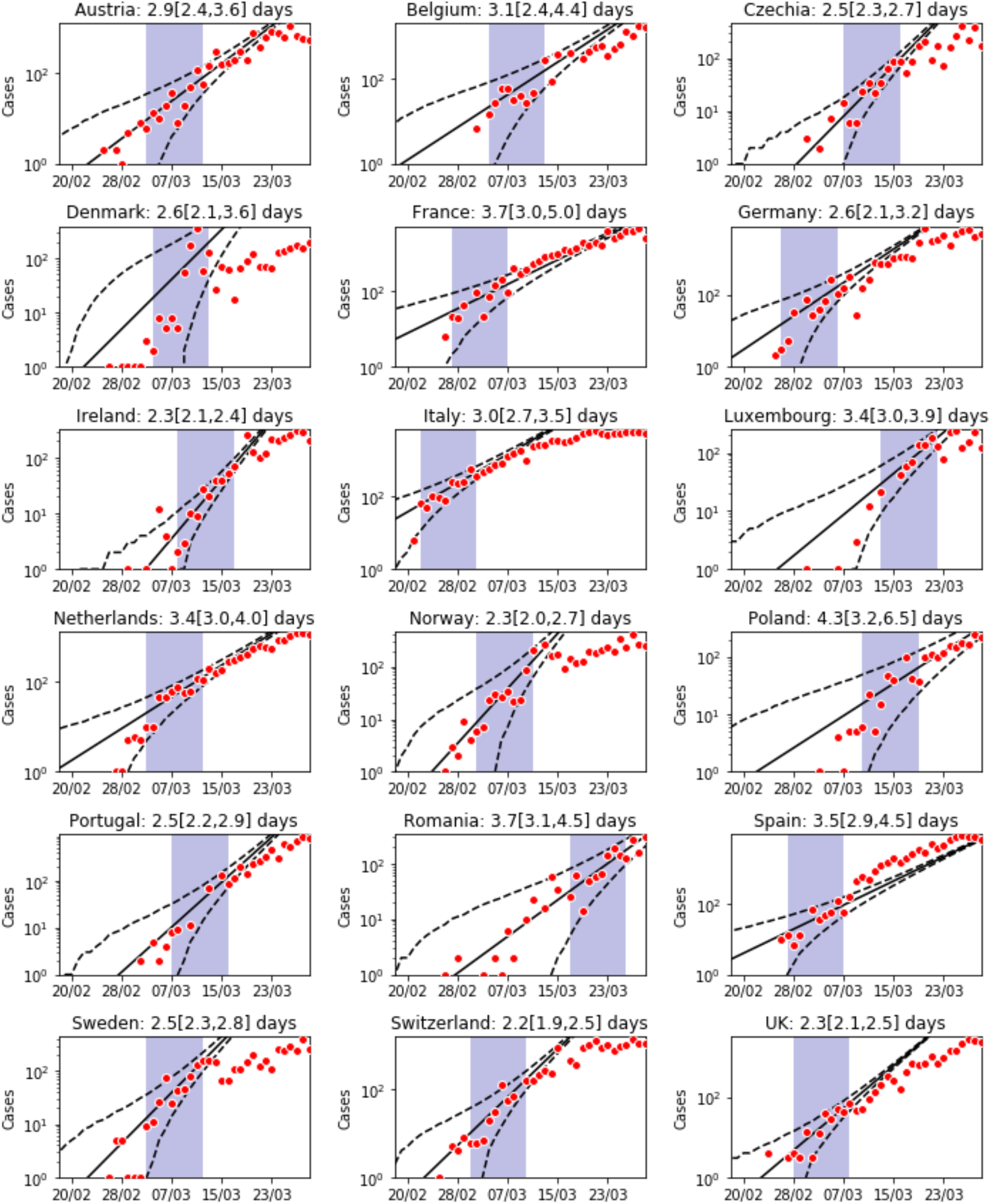
Unconstrained doubling times across Europe. Log daily confirmed cases (red dots) and estimated growth rate (solid) and 95% CIs (dashed black lines) for all European countries with more than 1000 cumulative confirmed cases by 27 March, obtained using a Generalised Linear Model (see Materials and Methods) in the 9-day data period after a cumulative incidence of 20 is reached (shaded area). Slight adjustments to Denmark and Romania reflect their particular circumstances (see Materials and Methods). Doubling times and 95% CIs are reported above each panel.

Although our results are robust to the method used, they might still be misleading if there are biases in the data. This issue is particularly critical as lags in reporting of cases can create discrepancies between national and international official sources (25-27) for case counts. However, these are unlikely to affect our conclusions owing to the following considerations:

1. The fast growth and high numbers likely make small biases negligible.
2. Any multiplicative correction, such as constant underreporting, does not affect the observed trend.
3. Exponential growth can easily be underestimated, e.g. if reporting rates decline over time, but is harder to consistently overestimate; aggressive swabbing of asymptomatic individuals (e.g. early on in the Italian locked-down towns), and changes in case definition, might introduce such a bias in the data, but are unlikely to affect observations for longer than a few days or consistently across different countries.
4. Hospital and ICU prevalence, which in the Italian data grow consistently with the number of confirmed cases (Figure 1B), are less affected by reporting issues. The observed faster increase in death rates may be explained by clustered outbreaks among vulnerable groups (e.g. care home residents), coupled with quicker progression to death among these groups, or possibly local hospital saturation.

Although each data stream individually has limitations, the evidence for fast exponential growth in the absence of intervention that emerges from their combination is compelling.

Because non-pharmaceutical interventions, with unknown adherence, cannot be evaluated until their effects emerge in the data, the delay between infection and case detection is crucial in determining how long cases have been growing (or, when lifting interventions, can grow) unobserved. Pre-symptomatic detection is not possible in the absence of full contact tracing and testing of asymptomatic individuals; detecting cases at symptom onset is potentially more feasible, but depends on the case-finding strategy, and frequently cases are not identified until hospital visit.

We estimated the incubation period and the delay between symptom onset and hospitalisation from UK line-list data provided by Public Health England (unfortunately not publicly available) and a publicly available line-list which collates worldwide data (28). Our results are more robust than previous published estimates, as we account for both truncated observations and exponential growth in the number of infected cases (see Materials and Methods), but nonetheless are consistent with the existing literature (Table 1). Our results also highlight geographical heterogeneity, such as shorter onset-to-hospitalisation intervals in Hong Kong and Singapore compared to the UK. With the exception of Singapore, the sum of the mean incubation period and mean onset-to-hospitalisation interval is never shorter than 9 days.

**Table 1.** Estimates of incubation period and delays from onset of symptoms to confirmation/ hospitalisation from the literature and from our estimation (see Materials and Methods).

| Parameter | Mean | Standard deviation | Confidence interval for the mean | Sample size | Source |
| --- | --- | --- | --- | --- | --- |
| Incubation period | 4.8 | - | 2.2 - 7.4 | 16 | (38) |
| Incubation period | 5.6 | 3.9 | 4.4 - 7.4 | 52 | (39) |
| Incubation period | 6.4 | 2.3 | 5.6 - 7.7 | 88 | (40) |
| Incubation period | 4.84 | 2.79 | - | 162 | Materials and Methods (Data - 28) |
| Onset to confirmation | 4.8 | 3.03 | - | 38 | (41) |
| Onset to hospitalisation | 5 | - | - | - | (42) |
| Onset to hospitalisation (dead) | 6.6 | - | 5.2 - 8.8 | 34 | (39) |
| Onset to hospitalisation (alive) | 9.7 | - | 5.4 - 17 | 155 | (39) |
| Onset to hospital visit (UK) | 5.14 | 4.20 | - | 90 | Materials and Methods (Data - PHE) |
| Onset to hospital visit (Singapore) | 2.62 | 2.38 | - | 92 | Materials and Methods (Data - 28) |
| Onset to hospital visit (Hong Kong) | 4.41 | 4.63 | - | 52 | Materials and Methods (Data - 28) |

Figure 3 illustrates the delays between infection and detection in early interventions in the UK and Italy. For both countries, after a control measure is implemented, the number of daily confirmed cases sustains the pre-intervention exponential growth for about 9 days, before deviating from the predicted trajectory.

**Fig. 3.**
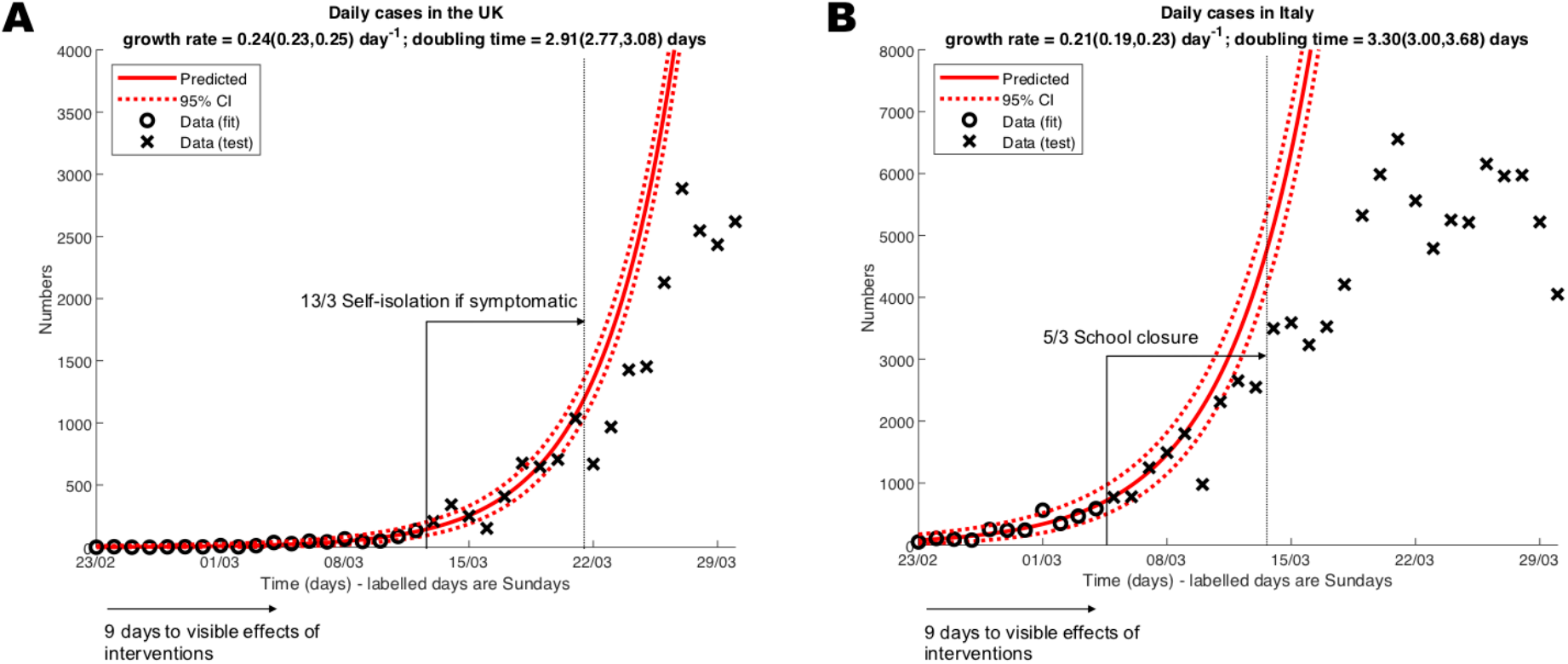
Visible delay of about 9 days between implementation of interventions and deviation from exponential growth. Daily confirmed cases in (A) the UK and (B) Italy before intervention (circles) are fitted with a Generalised Linear Model with 95% Negative Binomial CIs (dashed red lines; see Materials and Methods). Crosses are data not used for fitting.

Since *R*_0_ remains the mainstay of most epidemiological analyses, we determined values of *R*_0_ consistent with a range of growth rates and modelling assumptions. For a growth rate of 0.25/day and our estimates of the incubation period (Table 1), we obtained values ranging from 2 to 5 (Table S1A). This variability in *R*_0_ values at fixed growth rate is due to extreme sensitivity to modelling assumptions and parameters, in particular the extent of pre-symptomatic transmission (and hence of the generation time distribution), for which estimates in the literature vary widely (29). Conversely, the same *R*_0_ can be derived from different growth rates, if different generation times are assumed. For this reason, *R*_0_ values can be misleading if quoted in isolation: early estimates from China, which in the absence of reliable information were derived from relatively long SARS-based generation times, may be close to later estimates informed by shorter generation times, while hiding considerable differences in growth rates.

Moreover, with substantial pre-symptomatic transmission, the magnitude of *R*_0_ can correlate poorly with the feasibility of infection control (30). For example, if *R*_0_ = 4 and all transmission occurs after symptoms onset, self-isolation when symptomatic can achieve the required 75% fall in transmissions. Conversely, if *R*_0_ = 2 but most transmission is pre-symptomatic, the required 50% drop in transmissions can only be achieved through highly disruptive interventions, such as mass quarantine, unless a solid infrastructure is in place to rapidly detect and isolate pre-symptomatic individuals. Ultimately, neither *R*_0_ nor the growth rate can predict the precise impact of changes in physical distancing. However, unlike *R*_0_, growth rate and delays to detection can provide key guidance on how early action should be taken to successfully manage the risk that initial interventions, planned under significant uncertainty (e.g. in *R*_0_ values and expected levels of adherence), may later prove inadequate for halting growth.

## 3. Discussion

Our estimates of unconstrained doubling times from Europe provide rigorous reference values that can be used to remedy the paucity of these numbers in official sources. These values, which are twice as short as early estimates, are essential to plan worst-case scenarios and for calibrating models guiding the relaxation phase. Providing robust values for doubling time, delays, and *R*_0_, is vital to avoid reliance on outdated and potentially misleading estimates, and to avoid citation cascades which give them further prominence. For example, early figures from China still dominate the literature and government reports (31-34).

The combination of short doubling times and long delays between infection and case detection provides a powerful explanation for the pattern of countries initially underestimating the significance of a small number of observed cases but then, in the attempt to avoid the health-care system being overwhelmed, rapidly escalating interventions as each appears to have no effect. At unconstrained growth, even doubling local hospital capacity only buys three days of reprieve before bed capacity is breached. Moreover, even with immediate, severe interventions halting all community transmission, within-household transmission continues to occur, creating an additional delay between the beginning of interventions and their effect (35). Further delays in case-confirmations, hospitalisations, potential ICU admissions and deaths mean the latter figures continue growing well after transmission control has been achieved. By the end of March our early estimates were being used to advocate urgent action in countries still at the beginning of their epidemics (36). Today, they can support governments’ decisions for swift initial action and implementation of early and socially disruptive interventions, even when initial numbers appeared relatively low.

Recognition of both the growth rate and delays in case detection are fundamental to plan the successful easing of strong interventions. Growth rates provide a directly observable measure of current epidemic trends, whereas *R*_e_ needs to be estimated indirectly. Understanding of detection delays can avoid overconfidence in epidemic control: although likely not 8-fold as in the early phase, new infections might build up unnoticed for several days while detected cases appear consistently decreasing. Therefore, intervals between subsequent relaxations should be long enough to allow proper assessment of their impact and avoid the social challenges associated with re-introduction of stricter physical distancing. Significant levels of testing, coupled with standard and/or technology-based contact tracing, are essential to prevent transmission and shorten detection delays.

## Data Availability

All data and code used in this analysis are provided in the Github repository (https://github.com/thomasallanhouse/covid19-growth), with the exception of the UK onset-to-hospitalisation delay data. These data are provided by Public Health England under a data sharing agreement, and we are unable to share them.

## Acknowledgments

We acknowledge Alex Washburne and his collaborators, who at the time of initial submission were reaching similar conclusions from different directions, for valuable discussions. The initial motivation for this study comes from the open letter from Italy to the scientific community (37), but we thank the numerous members of the general public, especially from hard-hit countries, who have tried to voice these messages with whatever means they had.

## Author contributions

EB, IH, LP and FS collated epidemiological data. IH, TH, CO, and LP carried out the formal data analysis. KL, TH, LP, FS and HS were involved in the conceptualisation and development of ideas. All authors contributed to writing the manuscript.

## Ethical approval

No ethical approval was required for this work because the data used are either publicly available or do not allow individual subjects to be identified. Exemption was confirmed by the University of Manchester Ethics Decision Tool.

## Supplementary Materials

Details regarding the methods used in this analysis are provided in the supplementary material.

## Supplementary Materials for

**This PDF file includes:**

Materials and Methods Figs. S1 to S3 Table S1

**Other Supplementary Materials for this manuscript include the following:**

Data sources and the code used to carry out our data analysis can be found at: https://github.com/thomasallanhouse/covid19-growth

### Materials and Methods

#### Data Sources

We consider four sources for our epidemiological data: the WHO (*25*), line-list data provided by Public Health England (PHE), line-list data from (*28*) and the Italian Istituto Superiore di Sanità (*26*). Of these datasets, three are publicly available. The line-list from PHE is unfortunately not publicly available. From these sources, we extract epidemiological data concerning case counts, incidence, hospitalisation, and delays between infection and symptom onset, and onset of symptoms and hospitalisation.

These data sources and the code used to carry out our data analysis can be found at: https://github.com/thomasallanhouse/covid19-growth.

#### Fitting the growth rate

Typically, an infection spread from person to person will grow exponentially in the early phase of epidemic. This exponential growth can be measured through the real time growth rate *r* so that, loosely speaking, the incidence of infection is *y(t)* = *y_0_e^rt^* + noise. A natural mathematical model to derive the estimate of *r* is a Poisson family generalised linear model (GLM) with a log link.

The growth rate *r* is more intuitively reported as a doubling time (the time taken to double case numbers), defined as *t_D_* = ln(2)/*r*. If *r* < 0, then *t_D_* < 0 and it is often interpreted as halving time, *t_H_* = − *t_D_*. The log-linear analysis from a Poisson GLM defined formally below is restricted to datasets (or time windows) with clear exponential growth.

To allow, in semi-parametric manner, time variation in growth rates, we adapt a generalised additive model (GAM) where *y(t*) ∝ *e^s(t^*^)^ for some smoother *s(t*). Given the over-dispersed noise inherent in both disease dynamics and surveillance data, a quasi-Poisson family is considered here. A canonical link and a thin-plate spline is used as implemented in the R package *mgcv* (*43, 44*). The instantaneous local growth rate is then the time derivative of the smoother, *r_s_* = *ṡ(t*), and the instantaneous doubling time is calculated as *t_D_* = ln(2)/*ṡ(t*). Asymptotic confidence intervals are derived for *r_s_* which are only indicative of uncertainty on *t_D_*, especially as *r_s_* approaches zero and variance is relatively large.

Potential issues with the GAM approach include that extrapolation outside of the data range (and hence forecasting epidemic trend) is not sensible, and that there may be boundary effects from the choice of smoother. However, this approach has the major advantage that it allows for time-varying estimates of doubling time and thereby implicitly allows for missing explanatory information. We also include as additional explanatory variables a week day effect due to reduced case reporting at weekends, followed by a back-log of cases at the start of the week. Thus, denoting by *y(t*) the random number of new cases on day t, the expected value is

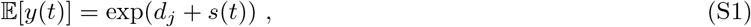

where *d_j_* takes a different value for each day of the week (*j* = 1, 2,…, 7) corresponding to time *t*.

Unlike the semi-parametric GAM approach, the parametric GLM approach lacks the ability to capture time variation, but allows for extrapolation and epidemiological interpretation. To capture over-dispersion we use a quasi-Poisson family for the noise model. Explicitly, the Negative Binomial probability mass function (pmf) is

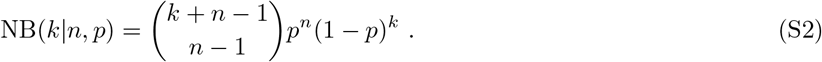

We will work in the parameterisation where the mean is *µ* and the variance is *θμ*, i.e.

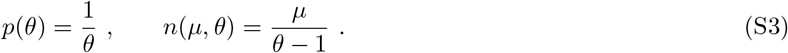

Let the number of new cases on day *t* be *y(t*). We assume that this is generated by an exponentially growing mean,

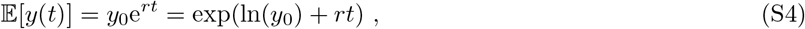

which is then combined with the negative binomial pmf to give likelihood function for observations over a set of times 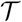 of

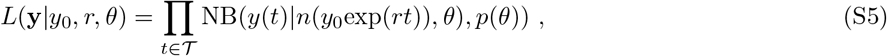

where 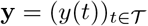. This can then be viewed as a GLM with time as a continuous covariate, intercept ln(*y*_0_), slope r, exponential link function and negative binomial noise model (*45*). We can perform inference through numerical maximum likelihood estimation (MLE) and calculate confidence intervals using the Laplace approximation (*46*).

Figure S1 shows the GAM fit on the number of daily confirmed cases reported by WHO (*25*) between 25 February and 31 March 2020 for all the 18 European countries that had reached 1000 cumulative confirmed cases by 27 March (the day Italy first overtook China). Data reported on day *t* + 1 are plotted against day *t*, assuming it takes one day for each country to report numbers to WHO, as is the case for Italy. For each country, the left panel shows the instantaneous growth rate from GAM (left axis) and the doubling time (right axis) with 95% CIs. The horizontal blue line shows a null growth rate, at which point the doubling time is infinitely long and below which the doubling time becomes negative. The right panel shows the output of the model fit and the data. For Belgium, the GAM selects as most parsimonious a model with no change in growth rate during the entire time interval. The apparently noisy data of Germany are explained well by a consistent day-of-the-week variability. Conversely, visible oscillations in the growth rate of Denmark, Norway and Sweden suggest this is not the case and reflect a more complex pattern in the data, likely due to wide variations in testing. This is also reflected in Figure S2, where the challenges in fitting a simple exponential growth are clearly visible.

#### Estimating delay distributions

Delay distributions describe the time delay between two events. To understand how long until the impact of an intervention may be observed, we need to understand the delay between infection and symptom onset (the incubation period), and the delay from onset to visiting hospital (or healthcare setting), since this is where cases are likely to be identified. A difficulty with estimating delay distributions during an outbreak is that events are only observed if they occur before the final sampling date. Since delay distributions depend on the time between two events, if the first event occurs near to the end of the sampling window, it will only be observed if the delay to the second event is short. This causes an over-expression of short delays towards the end of the sampling window, which is exacerbated by the exponential growth of the epidemic. Therefore, we need to account for this growth and truncation within our model.

To fit the data we use maximum likelihood estimation. However, we do not observe the delay directly, but rather the time of the two events. Therefore, we need to construct a likelihood function for observing these events. Following (*47*), we construct the conditional density function for observing the second event given the time of the first event and given that the second event occurs before date *T*. That is, we are interested in the conditional density function

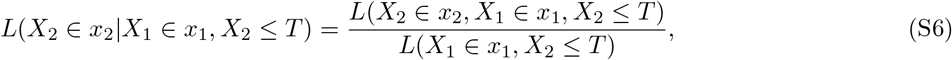

where *x*_1_ and *x*_2_ can be exactly observed or interval censored.

In the UK, the first healthcare visit is hospitalisation, as patients are encouraged not to seek in-person appointments with a general practitioner (GP) when showing flu-like symptoms. Therefore, we estimate the delay from onset to hospitalisation. This is estimated using FF100 data provided by Public Health England (unfortunately not publicly available), which contains data on the first few hundred infected individuals in the UK. These data incorporated the time of symptom onset and time of hospitalisation. Cases who were hospitalised before their onset date have been removed from the dataset, since they do not provide insight into the delay. Additionally, some cases have no symptom onset, so these have also been removed from the data. For cases where symptom onset and hospitalisation occur on the same day, we add half a day to the hospitalisation day, since the delay is unlikely to be instantaneous. After processing the data, this left 106 cases from which to infer the onset to hospitalisation delay. The dates in the line list are recorded exactly, so the likelihood function becomes

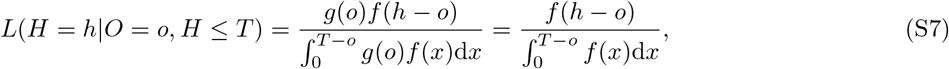

where *f* is the density of the onset to hospitalisation delay and *g* is the density of the onset time. Using this truncation corrected method and a gamma distribution to fit the delay distribution, we get a mean delay of 5.14 with standard deviation 4.20. To compare different regions, we also use data from Hong Kong and Singapore to estimate the local onset to healthcare visit delays. These data are taken from an open access line-list (*28*), and the filtered datasets are provided with the codes. Using the method above, for Hong Kong the mean delay is 4.41 days, with standard deviation 4.63, and for Singapore the mean delay is 2.62 with standard deviation 2.38. The mean delay is much shorter for Singapore because many of these healthcare visits occur in a clinic rather than hospital. Patients with mild symptoms attend a clinic, only going to hospital a few days later if their symptoms become significantly worse. Therefore, infected individuals will be identified faster, suggesting Singapore has a shorter observation delay. Hong-Kong data mostly refer to hospital admission, so the delay is similar to the UK.

For the incubation period, we use data from Wuhan during the early stages of the outbreak. These data were extracted from an open access line-list (*28*), containing dates when individuals were in Wuhan and when they developed symptoms (among other information). Since these data are from the early stages of the epidemic, the majority of cases were in Wuhan. Therefore, it is likely that these individuals were infected in Wuhan, so the time spent in Wuhan provides a potential exposure window during which infection occurred. For individuals with symptom onset date before leaving Wuhan or the same day they left Wuhan, the upper bound on the exposure window was adjusted to half a day before symptom onset. Using the data as of 21 February 2020, we have 162 cases from which to infer the incubation period. This infection date is interval censored, so we obtain the likelihood function

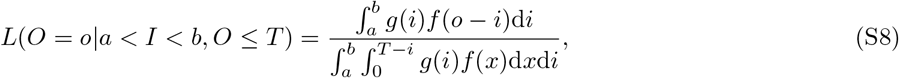

where *f* is the incubation period density function and g is the density function of the infection date. We assume *g* is proportional to the force of infection of the outbreak. Early estimates of the force of infection in Wuhan suggested a growth rate of around 0.1 (*9, 12*). However, these estimates were biased by stochasticity (high levels of noise in the dataset) in early identified cases before active case-finding was deployed, and the lack of testing and diagnosis guidelines at the time. More accurate estimates suggest that the early growth rate could be closer to 0.3 (*38*) or 0.4 (*48*). Based on our analysis of the European data and observations worldwide, the growth rate in the absence of interventions seems consistently between 0.2 and 0.3. Therefore, we assume the force of infection in Wuhan follows exponential growth with rate parameter 0.25 day^−1^. Using a gamma distribution to describe the incubation period, we obtain a mean incubation period of 4.84 days with standard deviation 2.79. Reducing the assumed force of infection to the early estimates (growth rate of 0.1 day^−1^) gives a mean incubation period of 5.92 with standard deviation 3.11. Therefore, although more accurate, using the growth rate of 0.25 does not substantially change the observation delay. Using the higher growth rate will more accurately capture the minimum length of the observation delay, which is what is important for the analysis in the main text.

#### Estimation of *R*_0_

The relationship between the growth rate r and the basic reproduction number *R*_0_ in a simple homogeneously mixing model is provided by the Lotka-Euler equation:

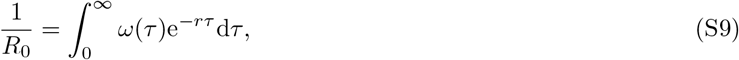

where *τ* represents the time since the infection of an individual and *ω*(*τ*) is the infectious contact interval distribution, defined as the probability density function (pdf) of the times (since infection) at which an infectious contact is made. An infectious contact is a contact that results in an infection if the contactee is susceptible, and early on in the epidemic any randomly selected contactee is almost surely susceptible.

Equation (S9) assumes all individuals have the same infectious contact interval distribution. However, if we assume random variability between individuals, there will be a set 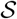 of curves *Ω*(*τ*). However, equation (S9) still applies, with *ω*(*τ*) being the time-point average of all curves in 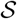 (*49*); see Figure S3.

The generation time *T_g_* is defined as the mean of the infectious contact interval distribution *ω*:

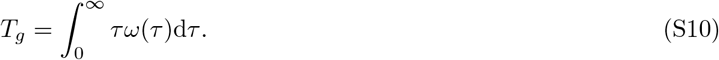

The same definition extends to a random infectivity profile of which ω is the time-point average.

For the incubation period we use our estimates from Table 1 (mean 4.84, standard deviation 2.79), which are anyway similar to those estimated by others. However, information about any form of pre-symptomatic transmission is hard to obtain but crucial for *R*_0_ estimates (*29*). Furthermore, there is also limited information concerning how infectivity changes over time. Therefore, Table S1 reports the estimates we obtain assuming the infectious period starts at the onset of symptoms, one, two or three days earlier, and assuming a Gamma-shaped infectivity with mean 2 or 3 days. In both cases, the standard deviation is assumed to be 1.5 and the infectivity is truncated after 7 days (see Figure S3).

We conclude that the estimates of *R*_0_ are highly sensitive to small variations in quantities that are challenging to estimate. However, for a growth rate of 0.25 day^−1^, close to what is observed in Italy and the UK, are also generally larger, and possibly much larger, than early official estimates (*5-8*). Smaller values in this range are associated with significant amounts of pre-symptomatic transmission, leading to a generation time compatible with some of the shortest estimates of the serial interval seen in the literature (*50*), and with a front-loaded infectivity curve (i.e. having mean 2, rather than 3).

We tested further assumptions. A simple SEIR model, with exponentially distributed incubation and infectious periods (with the same means as above but constant infectivity) leads to much smaller values of *R*_0_ than our estimates, as it favours very short incubation periods (Table S1B, left). Estimates, instead, do not change significantly if high variability in total infectiousness between individuals, in line with what observed for SARS, is assumed (Table S1B, right) or if 50% of cases are assumed to be fully asymptomatic and transmit at half the rate as those with symptoms (not shown).

These simple estimates are obtained under the assumption of mass-action mixing. The explicit presence of a social structure (e.g. age-stratification, household/network structure, etc.), which in principle could affect them, is likely negligible in such a high *R*_0_ and growth rate regime (*51*). The effect of the social structure on transmission is expected to grow in importance (especially the household structure, since isolation and quarantine facilitate within-household transmission) as the growth rate approaches zero.

**Fig. S1:**
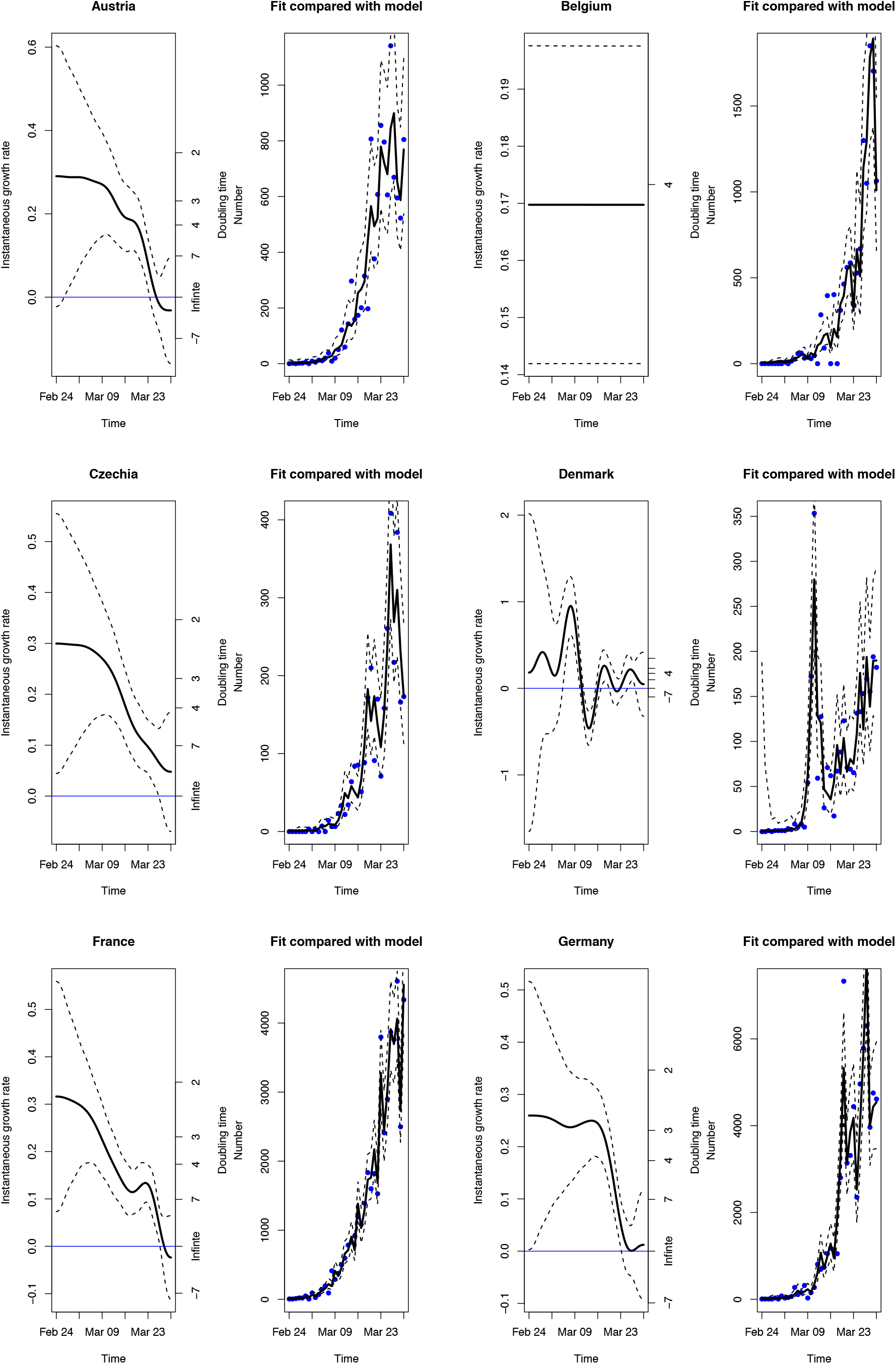
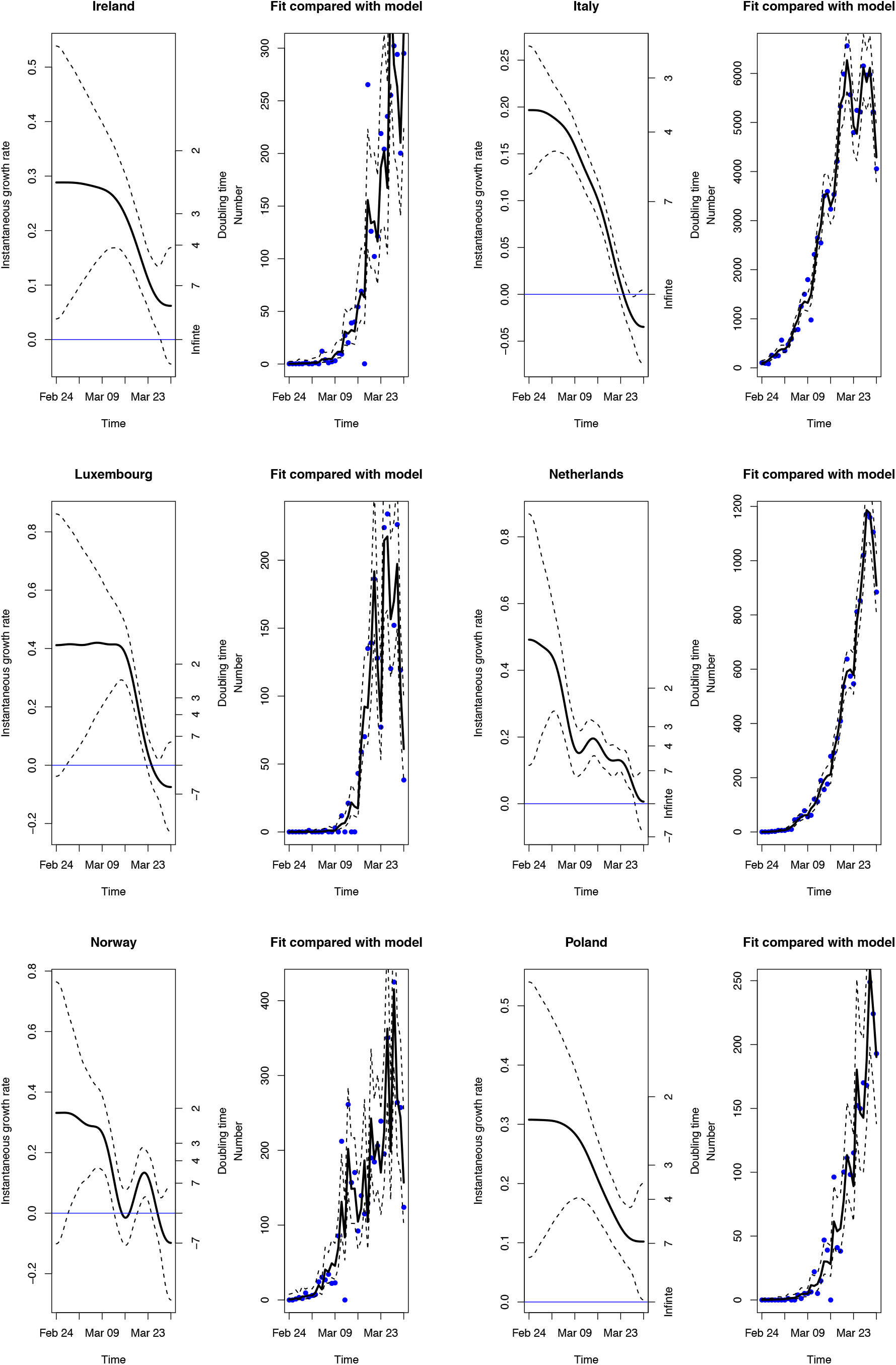
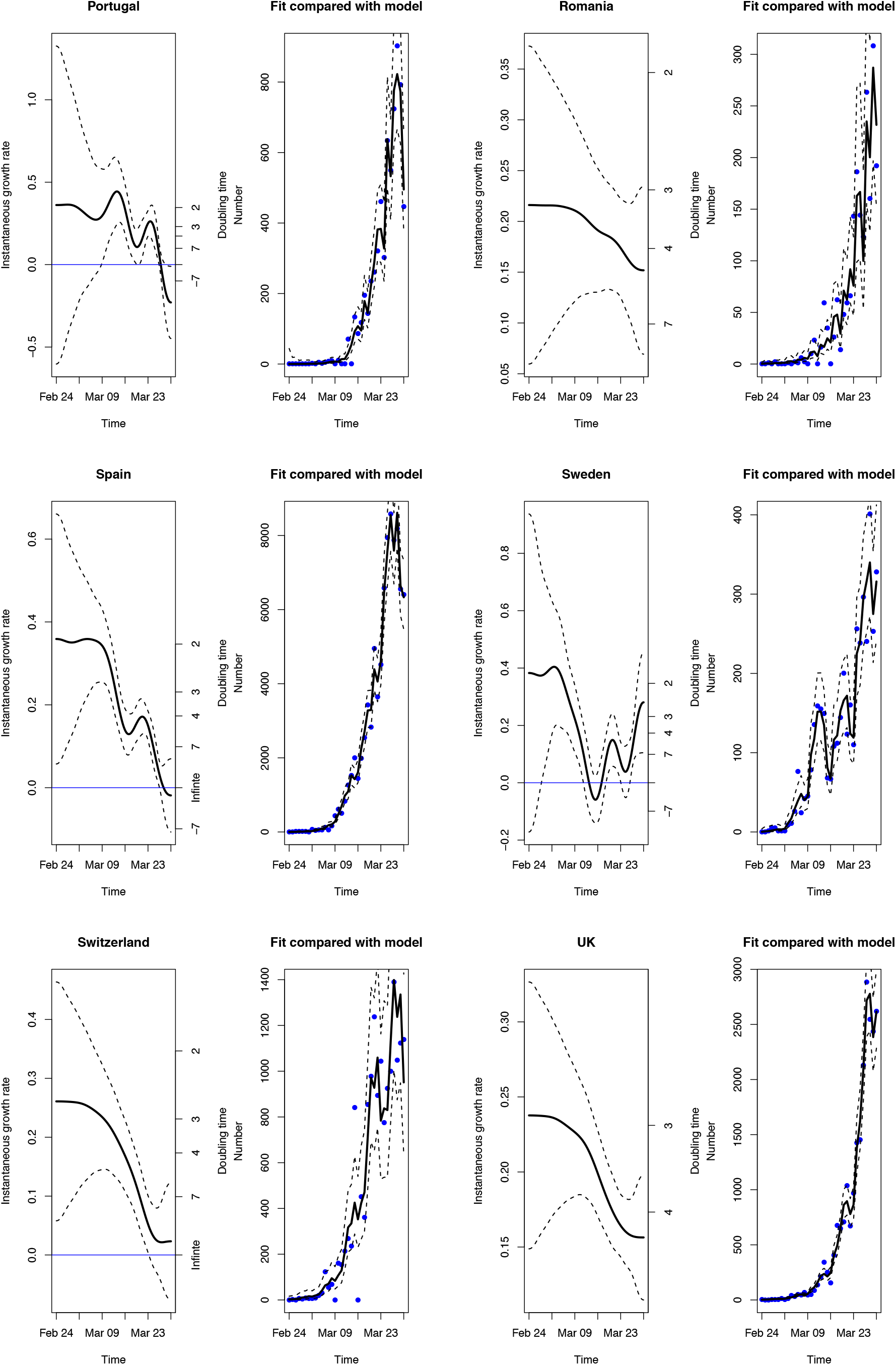
GAM fit for multiple European countries. Left panel: instantaneous growth rate and doubling time (solid line) with 95% CIs (dashed lines); right panel: model fit, which includes the day-of-the-week fixed effect, and data (blue dots).

**Fig. S2:**
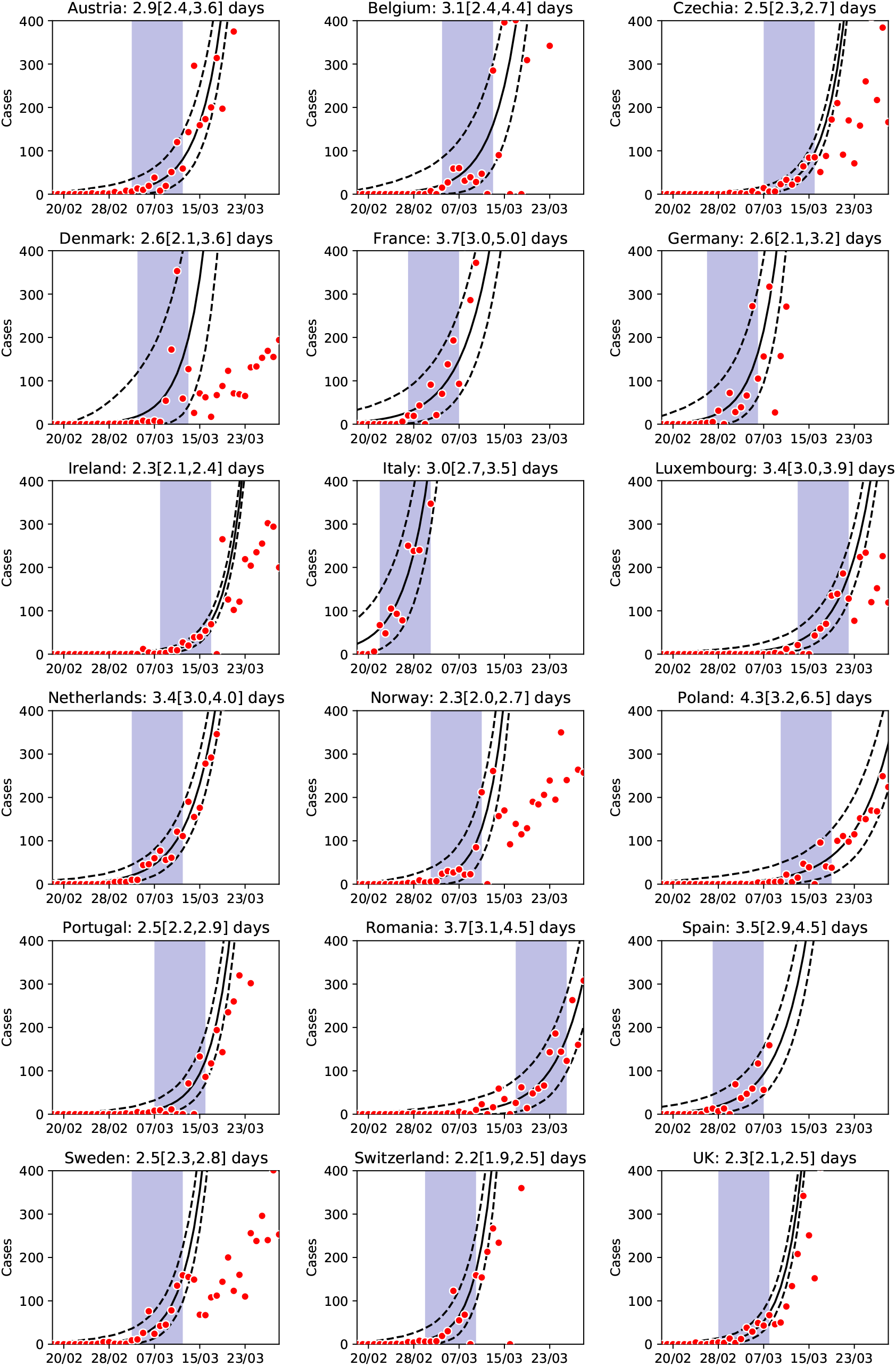
Linear *y*-axis version of Figure 2: daily confirmed cases (red dots) and estimated growth rate (solid) and 95% CIs (dashed black lines) for all European countries with more than a thousand cumulative confirmed cases by 27 March, obtained using a Generalised Linear Model in the 9-day data period after a cumulative incidence of 20 is reached (shaded area). The only exceptions are Romania, where the fit starts 7 days later to exclude a phase dominated by imported cases, and Denmark, where the fit starts 2 days earlier to capture the growth before the change in testing strategy.

**Fig. S3:**
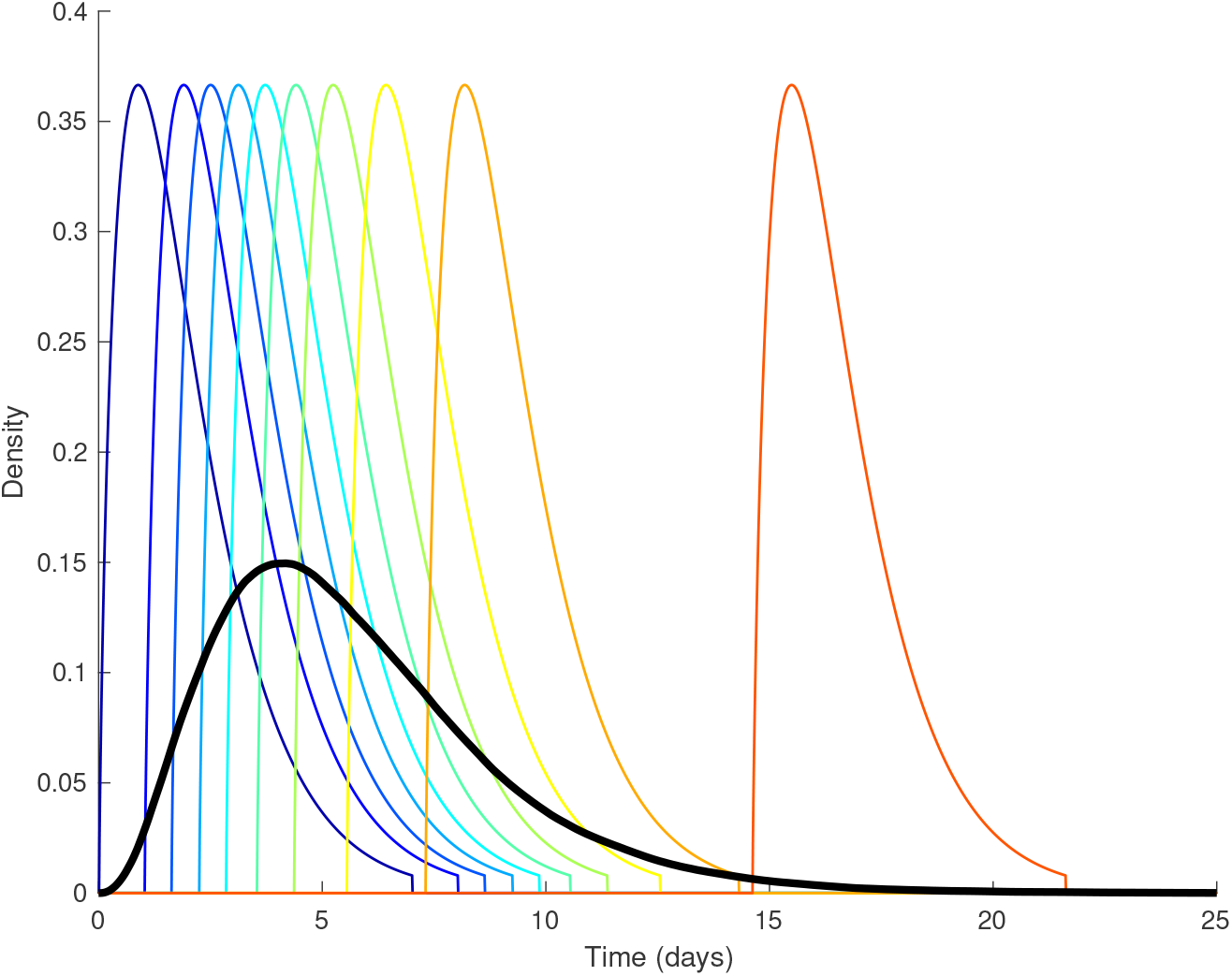
Example of random variability in the generation time distribution. The model assumes a Gamma-distributed latent period with estimates from Table 1, transmission starting 1 day before symptoms, a Gamma-shaped infectivity profile with mean 2 and standard deviation 1.5, and all individuals equally infectious. Ten profiles are drawn (thin coloured lines), out of the 10000 of which the thick black line is the time-point average.

**Table S1:** Values of *R*_0_ derived from different growth rates and different modelling assumptions. A) Gamma-distributed latent period with estimates from Table 1, and Gamma-shaped infectivity profile with mean 2 (left) and 3 (right) and standard deviation 1.5, assuming all individuals are equally infectious; B) SEIR model (left) and same model as in A (left) but assuming total infectiousness of each individual is randomly drawn from a Gamma distribution with mean 1 and standard deviation 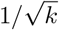, with k = 0.25.

| **A** |  | **$R_0$, assuming infectivity peaks in 2 days** |  |  |  | **$R_0$, assuming infectivity peaks in 3 days** |  |  |  |
| --- | --- | --- | --- | --- | --- | --- | --- | --- | --- |
| Growth rate<br>(1/day) | Doubling<br>time (day) | Days of pre-symptomatic transmission: |  |  |  | Days of pre-symptomatic transmission: |  |  |  |
|  |  | 0 | 1 | 2 | 3 | 0 | 1 | 2 | 3 |
| 0.1 | 6.93 | 1.88 | 1.70 | 1.55 | 1.40 | 2.03 | 1.84 | 1.66 | 1.52 |
| 0.15 | 4.62 | 2.51 | 2.16 | 1.87 | 1.62 | 2.81 | 2.42 | 2.09 | 1.83 |
| 0.2 | 3.47 | 3.29 | 2.71 | 2.24 | 1.85 | 3.81 | 3.14 | 2.58 | 2.16 |
| 0.25 | 2.77 | 4.25 | 3.34 | 2.64 | 2.10 | 5.09 | 4.00 | 3.15 | 2.54 |
| 0.3 | 2.31 | 5.42 | 4.07 | 3.09 | 2.36 | 6.70 | 5.04 | 3.80 | 2.95 |
| Generation time: |  | 6.76 | 5.76 | 4.78 | 3.72 | 7.56 | 6.55 | 5.51 | 4.59 |

| **B** |  | **$R_0$ for SEIR model** |  |  |  | **$R_0$ assuming variability in total infectiousness between cases** |  |  |  |
| --- | --- | --- | --- | --- | --- | --- | --- | --- | --- |
| Growth rate<br>(1/day) | Doubling<br>time (day) | Days of pre-symptomatic transmission: |  |  |  | Days of pre-symptomatic transmission: |  |  |  |
|  |  | 0 | 1 | 2 | 3 | 0 | 1 | 2 | 3 |
| 0.1 | 6.93 | 1.63 | 1.52 | 1.41 | 1.31 | 1.88 | 1.71 | 1.55 | 1.41 |
| 0.15 | 4.62 | 2.00 | 1.83 | 1.66 | 1.49 | 2.51 | 2.17 | 1.88 | 1.64 |
| 0.2 | 3.47 | 2.40 | 2.16 | 1.92 | 1.67 | 3.29 | 2.72 | 2.25 | 1.88 |
| 0.25 | 2.77 | 2.81 | 2.50 | 2.19 | 1.87 | 4.26 | 3.36 | 2.66 | 2.14 |
| 0.3 | 2.31 | 3.25 | 2.86 | 2.47 | 2.08 | 5.43 | 4.10 | 3.11 | 2.41 |
| Generation time: |  | 6.20 | 5.37 | 4.40 | 3.39 | 6.81 | 5.84 | 4.78 | 3.80 |

